# Glaucoma medication adherence and associated factors among adult glaucoma patients in Ethiopia: A systematic review and meta-analysis

**DOI:** 10.1101/2023.10.05.23296590

**Authors:** Kibruyisfaw Weldeab Abore, Estifanos Bekele Fole, Mahlet Tesfaye Abebe, Natnael Fikadu Tekle, Robel Bayou Tilahun, Fraol Daba Chinkey, Michael Teklehaimanot Abera

**Affiliations:** Department of Pediatrics, Yirgalem Hospital Medical College, Yirgalem, Sidama, Ethiopia; Department of Medicine, Yirgalem Hospital Medical College, Yirgalem, Sidama, Ethiopia; Long term care department, Burjeel Medical City, Abu Dhabi, United Arab Emirates; Bin Haider Healthcare Center, Dubai, United Arab Emirates; Department of radiology, College of Health Sciences, Addis Ababa University, Addis Ababa, Ethiopia

## Abstract

**Background:** Glaucoma medications are one important pillar of glaucoma management to control intraocular pressure. If left uncontrolled, intraocular pressure causes progressive visual loss and blindness. Thus, adherence to glaucoma medication is vital to prevent optic nerve damage and its consequences. This study was conducted to systematically summarize the magnitude of glaucoma medication adherence and the associated factors among adults with glaucoma in Ethiopia.

**Methods:** Database Searches to identify research articles was conducted on PubMed, EMBASE, Cochrane, AJOL, SCOPUS, and Google scholar without restriction on the date of publication. Data extraction was done using a data extraction Excel sheet. Analysis was performed using STATA version 16. Heterogeneity was assessed using I^2^ statistics. Pooled prevalence and pooled odds ratio with a 95% confidence interval using a random effect model were computed.

**Result:** We included 6 studies with a total of 2101 participants for the meta-analysis. The magnitude of adherence to glaucoma medication was found to be 49.46% (95% CI [41.27-57.66]). Urban residence (OR=1.89, 95% CI; 1.29-2.49), normal visual acuity (OR=2.82, 95% CI; 0.85-4.80, P=0.01), and payment means for medication (OR=0.22, 95% CI; 0.09-0.34) were found to be statistically significant predictors of adherence.

**Conclusion:** The magnitude of glaucoma medication adherence is lower than expected. Place of residence, visual acuity, and payment means had statistically significant associations with glaucoma medication adherence. Tailored health education on medication adherence and subsidization of glaucoma medication is recommended.

## Introduction

Glaucoma is a preventable cause of blindness which primarily is the disease of the optic nerve [1]. Globally, it is estimated to affect around 80 million individuals and cause blindness in 3.6 million individuals [2, 3]. Previous studies have shown that there are various risk factors to develop glaucoma. High intraocular pressure is a modifiable risk factor which is a target of treatment among glaucoma patients [4]. Currently available treatment modalities that include medical, surgical, and laser therapy are all primarily designed to lower the IOP [5].

Successful medical treatment of patients with glaucoma is contingent upon the adherence of the patients to the prescribed medication [6, 7]. Adherence is defined as the degree to which the patient takes the prescribed medication according to the prescription given by a health professional [6]. Adherence to glaucoma medication has been shown to decrease the progressive visual field loss and subsequent loss of vision among glaucoma patients [8, 9].

Although it is highly unreliable and prone to bias, self-reporting is the most commonly used method of assessing adherence [10, 11]. Previous studies done in different parts of the world have reported a highly variable level of non-adherence ranging from 5% to 80% [12]. Furthermore, Studies done in Ethiopia have also reported level of adherence ranging from 32.5% [13] to 61.4% [14]. Some of the factors associated with adherence to glaucoma medication include sociodemographic factors, poor understanding of the disease and its course, baseline line clinical condition, and medication related factors such as dosing, side effects, and costs of medication [15–19].

The magnitude of adherence and the factors affecting it reported in studies conducted in Ethiopia varies greatly. Therefore, this study aims to synthesize the pooled overall magnitude of glaucoma medication adherence and its associated factors among adult glaucoma patients in Ethiopia.

## Methods and materials

### Study design and Study setting

A systematic review and meta-analysis were done to assess the magnitude of glaucoma medication adherence and associated factors among adult glaucoma patients in Ethiopia. The protocol for the systematic review was registered on PROSPERO (Registration number: CRD42023449004).

### Search strategy

A comprehensive and systematic search of articles from PubMed, EMBASE, Cochrane, AJOL, SCOPUS, and Google scholar databases was made using the search terms ((“anti-glaucoma” OR “glaucoma medication” OR “glaucoma drug” OR “topical anti-glaucoma”) AND (adherence OR compliance)) AND (“Ethiopia”). The full PubMed search was (“anti-glaucoma”[All Fields] OR “glaucoma medication”[All Fields] OR “glaucoma drug”[All Fields] OR “topical anti-glaucoma”[All Fields]) AND (“adherance”[All Fields] OR “adhere”[All Fields] OR “adhered”[All Fields] OR “adherence”[All Fields] OR “adherences”[All Fields] OR “adherent”[All Fields] OR “adherents”[All Fields] OR “adherer”[All Fields] OR “adherers”[All Fields] OR “adheres”[All Fields] OR “adhering”[All Fields] OR (“compliances”[All Fields] OR “patient compliance”[MeSH Terms] OR (“patient”[All Fields] AND “compliance”[All Fields]) OR “patient compliance”[All Fields] OR “compliance”[All Fields] OR “compliance”[MeSH Terms])) AND “Ethiopia”[All Fields]. The search was made up to September 2023 with restriction to only articles published in English. The Preferred Reporting Items for Systematic reviews and Meta-Analyses (PRISMA) guideline [20] was used to document and report the steps of study selection.

### Eligibility criteria

#### Inclusion criteria

This review included studies (cross-sectional, cohort, and case-control studies) that evaluated glaucoma medication adherence and/or its associated factors among adults in Ethiopia with no restriction on the year of publication of the study.

#### Exclusion criteria

Studies conducted outside of Ethiopia and studies which are not published in English language were excluded from the study.

### Quality appraisal

Quality assessment of the included studies was done using the Newcastle-Ottawa scale (NOS) by three reviewers independently [21]. It assesses bias in 3 domains namely selection, comparability, and outcome domains. Those with a grade of less than 7 were assessed as poor and those with greater than or equal to 7 were assessed as having good quality. Disagreement in scoring between reviewers was settled by discussion with the help of a fourth reviewer.

### Data extraction

Two reviewers searched databases and applied the eligibility criteria independently to select studies. Covidence was used to manage articles. Examination of the selected studies against the eligibility criteria was done by two reviewers. Data extraction from selected studies was done by two reviewers independently and examination of the extracted data was done by a third reviewer. Differences between the reviewers were resolved through discussion and when agreement was not reached a third reviewer was involved to mediate. A pre-prepared EXCEL data extraction sheet was utilized to extract data from included studies which included the name of the primary author, region of the country, year of publication, study design, sample size, magnitude of adherence to anti-glaucoma medication, and risk factors with the corresponding measure of effect.

### Data analysis

Data was exported from Excel to STATA v.16 for analysis. Pooled magnitude of adherence and pooled odds ratio with a 95% confidence interval was computed using random effect model with the DerSimonian-Laird method and presented using a forest plot. To calculate the pooled odds ratio, we included variables which were reported as a statistically significant variable in at least two studies. Heterogeneity was assessed using I^2^ statistics and meta-regression was done to identify potential sources of heterogeneity. Furthermore, subgroup analysis was conducted based on the method of adherence ascertainment used by the studies. Publication bias was assessed using funnel plot and Eggers test.

## Result

### Search results

After searching various databases, we were able to retrieve a total of 172 articles. We removed 20 articles due to duplication and 144 articles were excluded after reviewing the title and abstract. After a full article review, 6 studies reporting wrong outcomes were excluded. In addition, 1 study with a full article not retrievable and 1 study done in a similar hospital as the study included in the final review were excluded from the review. A total of 6 Articles were considered for full article review against the eligibility criteria and included in the final review. (Fig 1)

**Fig 1.**
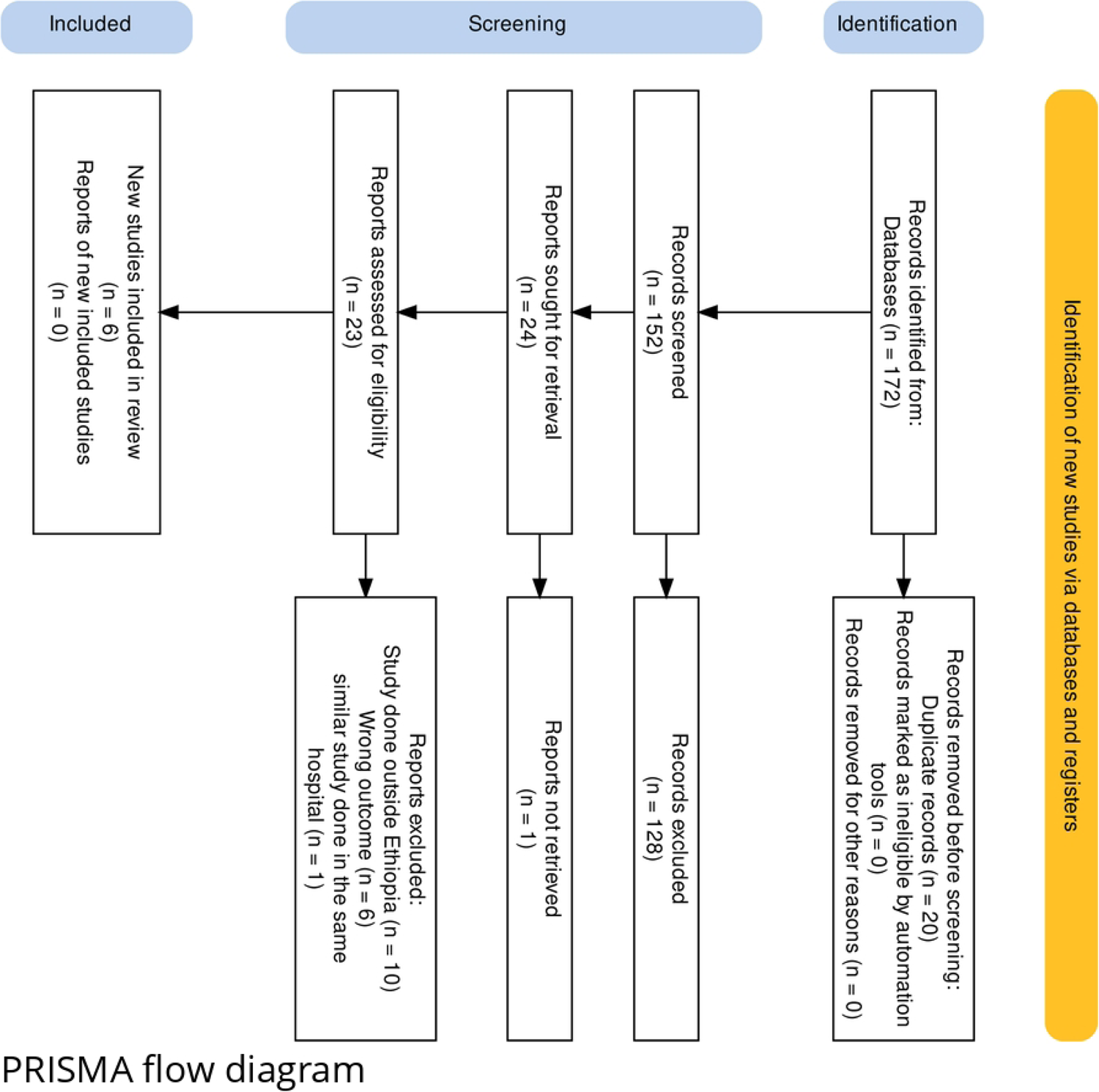
PRISMA study selection flow for systematic review and meta-analysis of Glaucoma medication adherence and associated factors among adults in Ethiopia.

### Study characteristics

All of the included primary studies used a cross-sectional study design. Of the studies, 1 was done in Sidama region [22], 1 in Addis Ababa [23], 1 study done in Oromia region [13], and 3 studies were done in Amhara region [14, 24, 25]. The studies were published between 2015 and 2023 and included a sample size ranging from 200 [13] to 410 [22]individuals. The total number of participants included in the review was 2101. Regarding the method of ascertainment of adherence, all of the included studies used self-reporting by the patient to declare adherence. Of the studies, 4 of them used a tool designed by the authors for the study [13, 14, 22, 24] while 2 studies used the Morisky medication adherence scale-8 [23, 25]. All included studies had good quality based on NOS score.

**Table 1.** Study characteristics of included studies on Glaucoma medication adherence and associated factors among adult glaucoma patients in Ethiopia.

| Authors | Year of publication | Region | Study design | SS | NOS score | Adherence (95% CI) |
| --- | --- | --- | --- | --- | --- | --- |
| Melaku M. Et al[22] | 2023 | Sidama | Cross-sectional | 410 | 7 | 53.9% (48.8-58.5) |
| Birhanie et al[25] | 2022 | Amhara | Cross-sectional | 382 | 8 | 49.5% (44.4-54.5) |
| Assem et al[24] | 2020 | Amhara | Cross-sectional | 390 | 7 | 56.2% (51-62) |
| Anbesse et al[14] | 2018 | Amhara | Cross-sectional | 360 | 7 | 61.4% (56.1-66.7) |
| Mehari T. et al[23] | 2016 | Addis Ababa | Cross-sectional | 359 | 7 | 42.6% (37.38-47.83) |
| Tamrat et al[13] | 2015 | Oromia | Cross-sectional | 200 | 7 | 32.5%(25.8-39.2) |

SS; sample size, NOS; Newcastle Ottawa score

### Magnitude of glaucoma medication adherence

We used 6 studies with a total of 2101 participants to estimate the pooled prevalence of magnitude of glaucoma medication adherence. There was a statistically significant high heterogeneity among studies based on I^2^ (I^2^=92.82%) and a random effect model with the DerSimonian-Laird method was used to determine the pooled magnitude. The magnitude of adherence among adult glaucoma patients was found to be 49.46% (95% CI; 41.27-57.66) (**Fig 2**). To explore potential sources of heterogeneity, meta-regression was done using sample size and publication year as covariates and both had no effect on the heterogeneity (**Table 2**). Sub-group analysis was done based on the method used to classify adherence. The pooled magnitude of adherence for studies that used a tool prepared by the author was found to be 51.13% (95% CI; 38.83-63.43) and for studies that used the MMAS-8 it was found to be 46.08% (95% CI; 39.32-52.85) (**Fig 3**).

**Fig 2.**
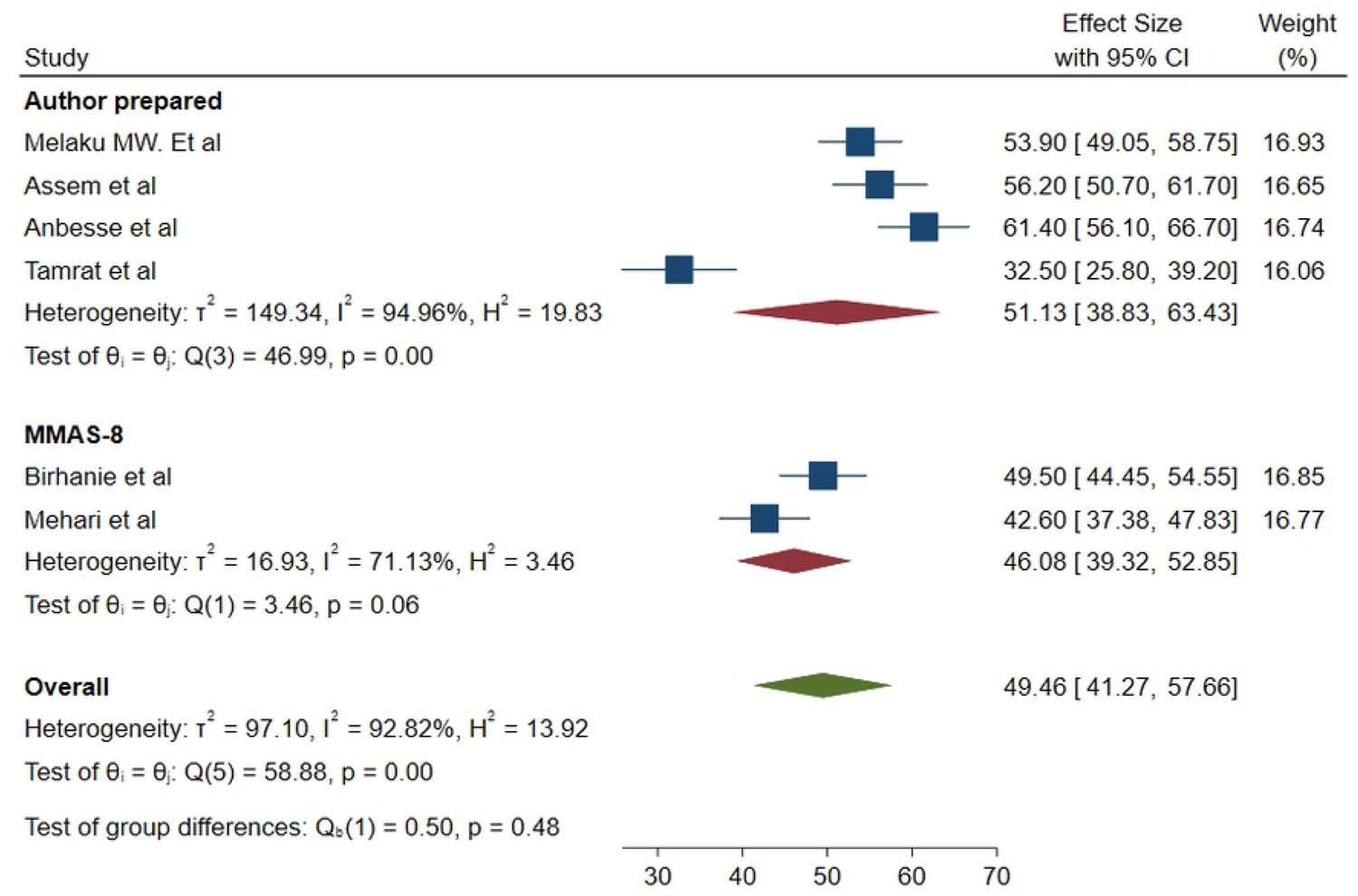
Forrest plot for the pooled magnitude of adherence to glaucoma medication among adult glaucoma patients in Ethiopia.

**Fig 3.**
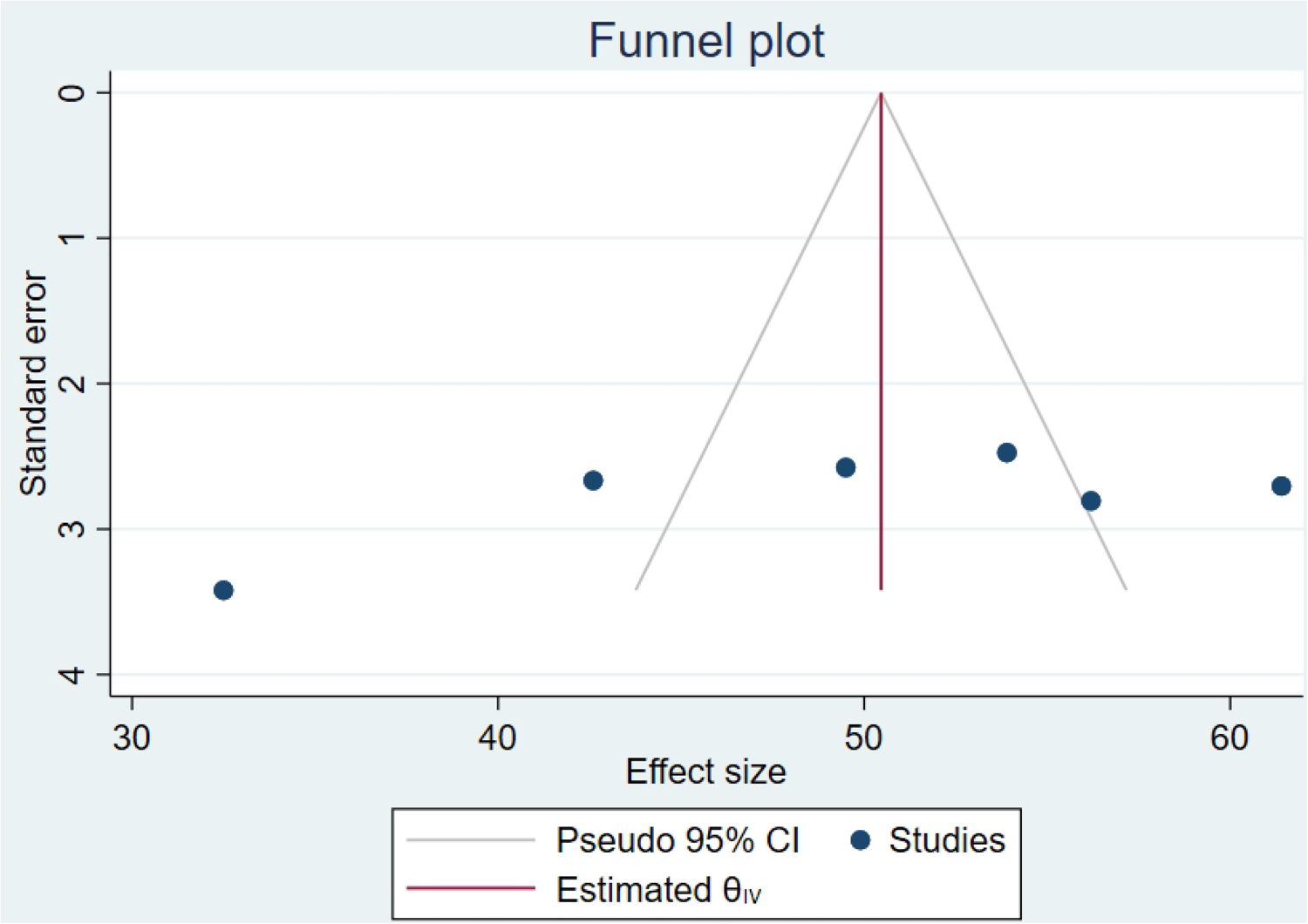
Forrest plot for the pooled magnitude of adherence to glaucoma medication among adult glaucoma patients based on the method of adherence classification used.

**Table 2.** Meta-regression analysis of factors affecting between-study heterogeneity.

| Source | Coefficient | Std. error | p-value |
| --- | --- | --- | --- |
| Sample size | .109635 | 0.076137 | 0.15 |
| Publication year | -0.04151 | 1.763183 | 0.981 |

### Publication bias

Publication bias was assessed using a funnel plot and Eggers test (p=0.0596) which revealed there was no publication bias (**Fig 4).**

**Fig 4.**
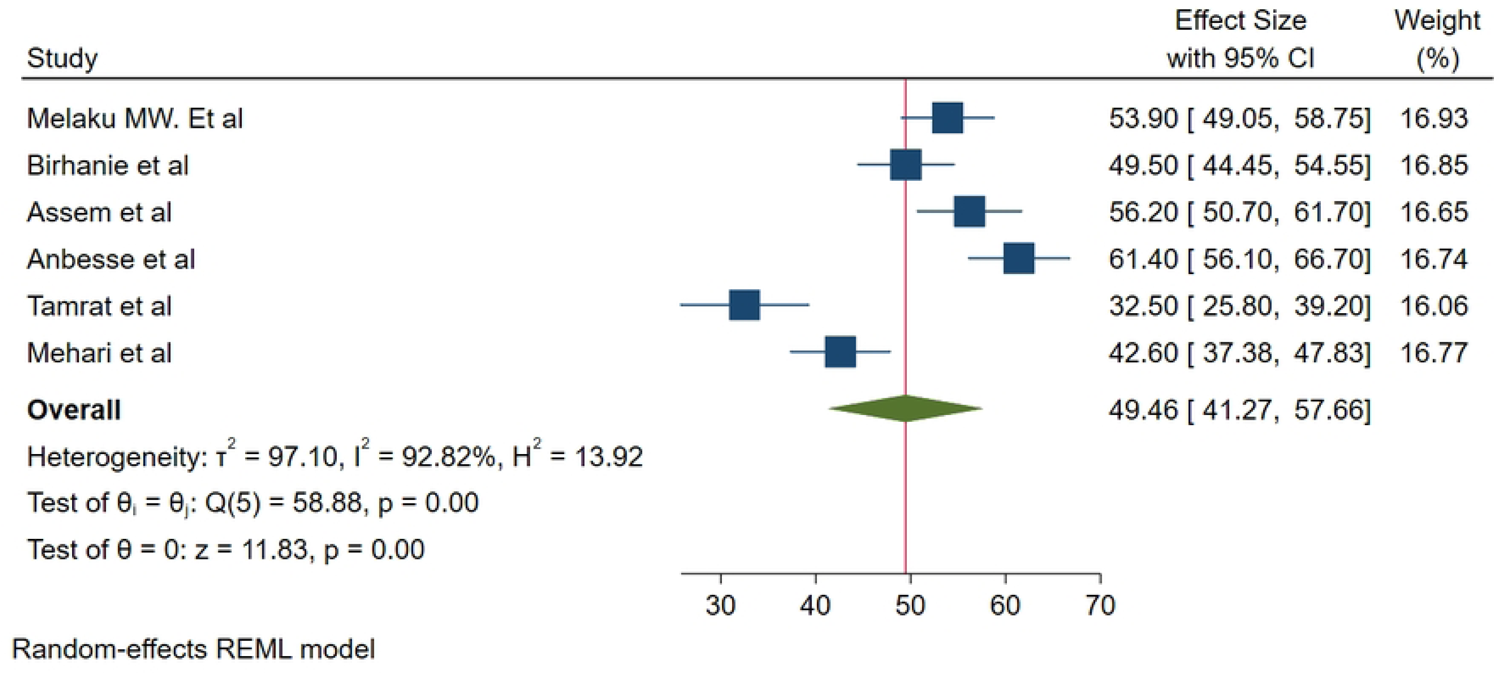
Funnel plot assessing publication bias of the magnitude of glaucoma medication adherence among adult glaucoma patients in Ethiopia.

### Factors associated with glaucoma medication adherence

We included variables that were significantly associated with adherence in at least two or more of the studies.

### Association between place of residence and glaucoma medication adherence

We used three studies with 1160 participants [14, 22, 24] to assess the association between place of residence and adherence. The odds of adherence to glaucoma medication was found to be 1.89 times higher among urban residents than rural residents (OR=1.89, 95% CI; 1.29-2.49). (**Fig 5)**

**Fig 5.**
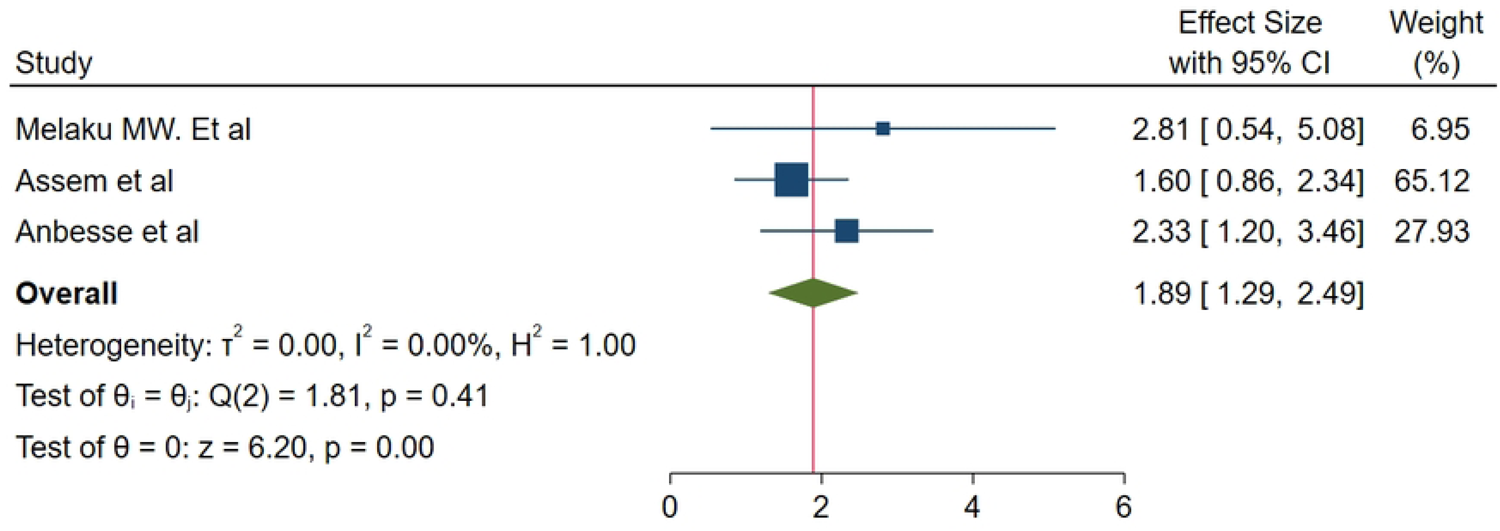
Association between place of residence and glaucoma medication adherence.

### Association between visual acuity and glaucoma medication adherence

We used three studies with 1160 participants [14, 22, 24] to assess the association between visual acuity and adherence. The odds of adherence to glaucoma medication was found to be 2.82 times higher among patients with normal visual acuity than those with affected visual acuity (OR=2.82, 95% CI; 0.85-4.80, P=0.01). (**Fig 6)**

**Fig 6.**
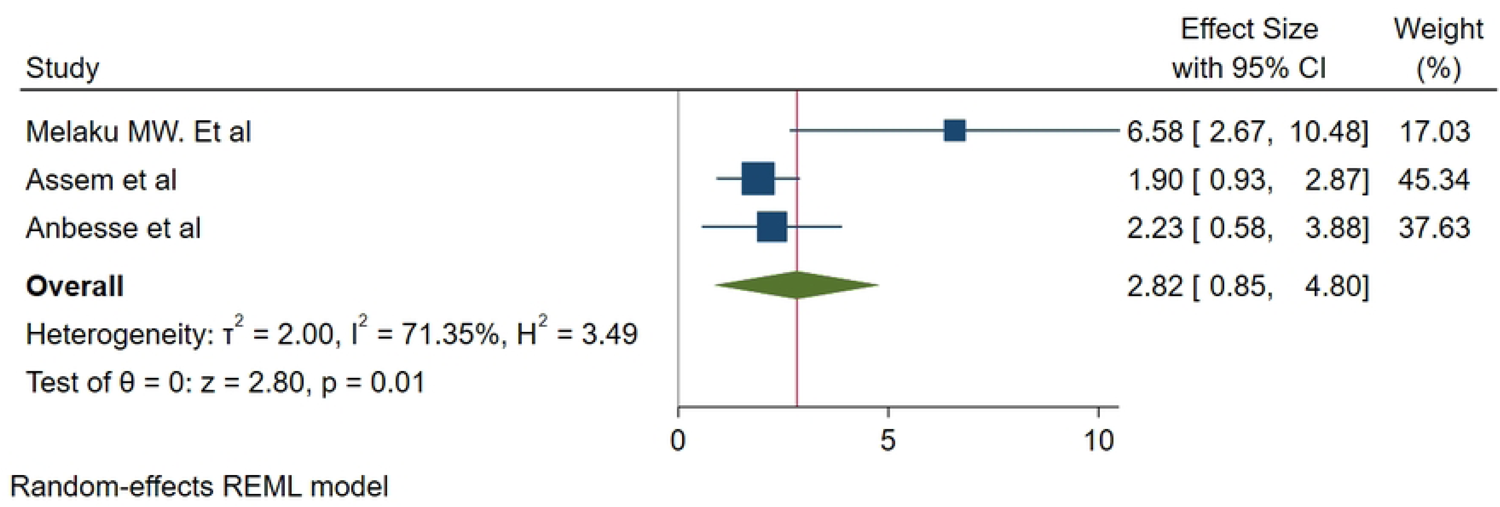
Association between visual acuity and glaucoma medication adherence.

### Association between means of payment and glaucoma medication adherence

We used two studies to examine the association between means of payment for medication and adherence with a total of 719 study participants [14, 23]. It was found that those who pay for the glaucoma medication themselves had 78% lower odds of adherence to glaucoma medication than those who are sponsored (OR=0.22, 95%CI; 0.09-0.34). (**Fig 7)**

**Fig 7.**
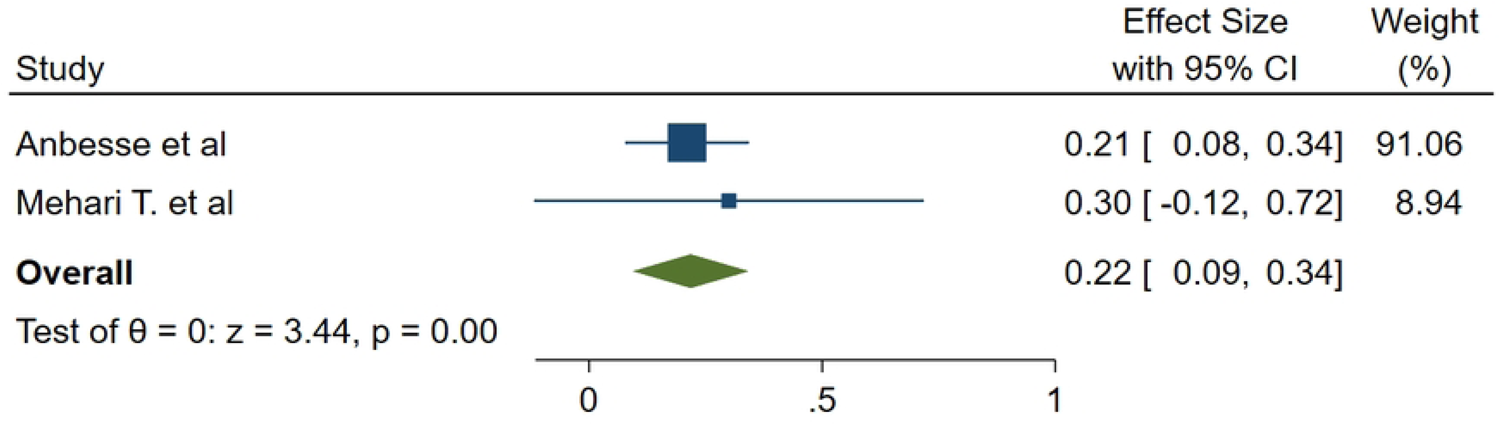
Association between means of payment and glaucoma medication adherence.

## Discussion

A systematic review and meta-analysis were done to determine the magnitude of glaucoma medication adherence and factors associated with it among adult glaucoma patients. It was found that less than half of glaucoma patients (49.46%) adhered to the prescribed medication. This finding is comparable with a study done in Ghana (48.5%) [26], Egypt (46.4%) [27], and higher than reported in Togo (10%) [28] and Iran (34.6%) [17]. However, it is lower than studies conducted in Benin (53%) [29], Israel (71%)[30], Saudi Arabia (72.6%) [31], and Korea (72.6%)[32]. This variation could be due to differences in socioeconomic profiles, differences in the health systems and health insurance coverage of the countries, differences in definition and method of ascertainment of adherence as well as variation in sample size.

In this study, it was found that those residing in urban areas had 1.89 times higher odds of adherence than those residing in rural areas. A study done in Iran in 2015 had also reported a statistically significant association between place of residence and medication adherence [17]. This could be due to the differences in access to health care and health related information and differences in access to health services like pharmacies for medication refill.

Furthermore, those with normal vision had 2.82 times higher odds of adherence to glaucoma medication compared to those with affected vision. This could be related to the dissatisfaction with the lack of change in the disease among those with affected visual acuity [17, 30]. Additionally, those who have impaired visual acuity are highly likely to be dependent for their day to day activities including application of the medication, purchasing medication, and going to health facilities for follow-up. A study done in Israel had also shown that there is a statistically significant association between functional dependence and glaucoma medication adherence (p=0.005)[30].

Those who paid for the medication by themselves had 78% lower odds of adherence than those who were sponsored for their medication. This finding is in comparative agreement with studies done in Nigeria [33], Ghana [34], and Romania[35] that reported the cost of the medication as a significant barrier to glaucoma medication adherence. The finding could be attributed to the long course of treatment, the high cost of glaucoma medication coupled with the poor socioeconomic status of Ethiopian community. This effect could be more pronounced among those who are on multiple glaucoma medications [17, 27] and those with comorbidities taking other medications.

### Limitation

This study is affected by recall bias and desirability bias inherent to the primary studies that used a self-reporting method of adherence ascertainment. This could potentially lead to overestimation of adherence to medication. Moreover, only 4 out of the 12 regions of Ethiopia are represented in the study which makes the finding less generalizable to Ethiopia.

## Conclusion

The magnitude of adherence to glaucoma medication adherence is low among adult glaucoma patients in Ethiopia. Area of residence, visual acuity, and means of payment had a statistically significant association with glaucoma medication adherence. Tailored health education to those coming from rural areas and those with affected visual acuity regarding glaucoma and drug adherence is recommended. Additionally, availing glaucoma medication at a subsidized price to those who cannot afford to buy medication and expanding community health insurance is recommended. We also recommend further studies using objective methods of assessment of adherence to be done.

## Data Availability

All relevant data are within the manuscript and its Supporting Information files

## Supporting information

**S1. PRISMA 2020 checklist**

**S2. Data set**

